# Multiscale heterogeneity of functional connectivity in autism

**DOI:** 10.1101/2024.10.20.24315248

**Authors:** Iva Ilioska, Marianne Oldehinkel, Alberto Llera, Maroš Rovný, Ting Mei, Seyed Mostafa Kia, Dorothea L. Floris, Julian Tillmann, Rosemary J. Holt, Eva Loth, Tony Charman, Declan G. M. Murphy, Christine Ecker, Tobias Banaschewski, Maarten Mennes, Christian F. Beckmann, Andre Marquand, Jan K. Buitelaar, Alex Fornito

## Abstract

Atypical functional connectivity (FC) in autism is a common finding, but the results of individual studies are often inconsistent and sometimes contradictory. Classical reliance on case-control comparisons of group means that ignore the inter-individual heterogeneity in autism may be a key drive of this inconsistency. Here, we used normative modelling to examine FC heterogeneity at the level of pair-wise inter-regional connections, specific brain regions, and broader functional networks in 1,824 participants (796 autistic) aged 5-58 years recruited across 32 different sites. Connection-level heterogeneity was high in both groups, with no single connection deviating in more than 4% of participants. However, deviant connections tended to converge on common regions and networks in autistic individuals more than in controls. Autistic individuals showed significantly greater overlap for positive deviations (i.e., atypically increased FC) in transmodal systems and negative deviations (atypically decreased FC) in sensory-motor areas. FC deviation patterns across coarser levels correlated with social functioning symptoms and intellectual ability. This work suggests that clinical variability in autism may be associated with extreme heterogeneity in the specific functional connections, whereas commonalities may be driven by convergence of atypical FC increases in transmodal systems and atypical decreases in sensorimotor networks, pointing to an imbalance in the functional organization of the brain’s sensorimotor-association axis.

## INTRODUCTION

Autism is a complex neurodevelopmental condition characterized by atypical social interaction and communication, restricted interests, repetitive behaviors, and atypical sensory processing (APA, 2013). Despite this core set of common symptoms, the underlying causes, biology, and outcomes of the disorder can vary greatly from person to person (King et al., 2019).

Heterogeneity is hypothesized to be a pervasive characteristic of autism, affecting every domain from behavioral manifestations to neural connectivity (Happé, Ronald, & Plomin, 2006; Jocelyn V Hull et al., 2017). This multifaceted heterogeneity poses a considerable challenge for replicating findings and discovering biomarkers (Lombardo, Lai, & Baron-Cohen, 2019; Masi, DeMayo, Glozier, & Guastella, 2017). Given this context of heterogeneity, it is perhaps unsurprising that despite autism being commonly viewed as a disorder of brain connectivity, decades of research have revealed largely inconsistent and contradictory findings with respect to the specific brain areas implicated and the direction (i.e., increases, decreases, or both) of group differences in functional connectivity (FC) being reported (Jocelyn V Hull et al., 2017). Although mega-analyses have identified robust, brain-wide FC differences between patients and controls (A. Di Martino et al., 2014; Holiga et al., 2019; Ilioska et al., 2023), the lack of reproducible, person-specific biomarkers means that a diagnosis of autism is still based solely on behavioral observation (American Psychiatric Association & Association, 2013).

One factor affecting our ability to parse the heterogeneity of autism and potentially identify reliable biomarkers is a reliance on traditional case-control comparisons of group means, which do not adequately characterize individual variability in neurobiological or clinical characteristics (Segal et al., 2024). The recent development of normative modeling has allowed researchers to move beyond mean-centric analyses to quantify how individual values for a given phenotype deviate from a model of normative expectations given a person’s age, sex, and other relevant demographic characteristics (Marquand et al., 2019; Marquand, Rezek, Buitelaar, & Beckmann, 2016). This approach has been used for neuroanatomical phenotypes in depression, bipolar disorder, schizophrenia, ADHD, and autism, and consistently identifies substantial heterogeneity between patients, indicating that the group mean may represent a poor summary of the effects of a particular diagnosis in individual patients (Marquand et al., 2019; Segal et al., 2022; Wolfers et al., 2020; Zabihi et al., 2020; Zabihi et al., 2019).

One normative modeling study of cortical thickness in autism reported highly individualized patterns of thickness deviations that differed across developmental stages in autism (Zabihi et al., 2019). Another study found that person-specific deviations of regional grey matter volume were rarely located in the same brain region, but that they were embedded within common functional circuits and networks across people, offering a putative neurobiological substrate for clinical differences (regional heterogeneity) and similarities (circuit- and network-level overlap) between autistic people (Segal et al., 2022).

Despite the widespread assertions about heterogeneity in autism, there has been a lack of comprehensive quantification and direct comparison of this variability regarding functional connectivity. Here, we applied a multiscale normative modeling approach to characterize and compare the inter-individual heterogeneity of FC in autism at three levels: single connections, regions, and within and between canonical functional networks. We hypothesized that single FC connections identified as deviant will show minimal overlap among individuals, suggesting a high degree of individual variability that may underlie the inconsistent findings frequently reported in the autism literature (Figure 1A). We further hypothesized that these deviations would aggregate within specific brain regions (Figure 1B) and show a preferential concentration within and between functional networks (Figure 1C), more consistently in autism. We then assessed the predictive accuracy of extreme deviations at the level of connections, regions, and networks in relation to clinical measures assessing the three primary symptom domains of autism—social difficulties, restricted and repetitive behavior, and sensory processing—in addition to full-scale IQ.

**Figure 1.**
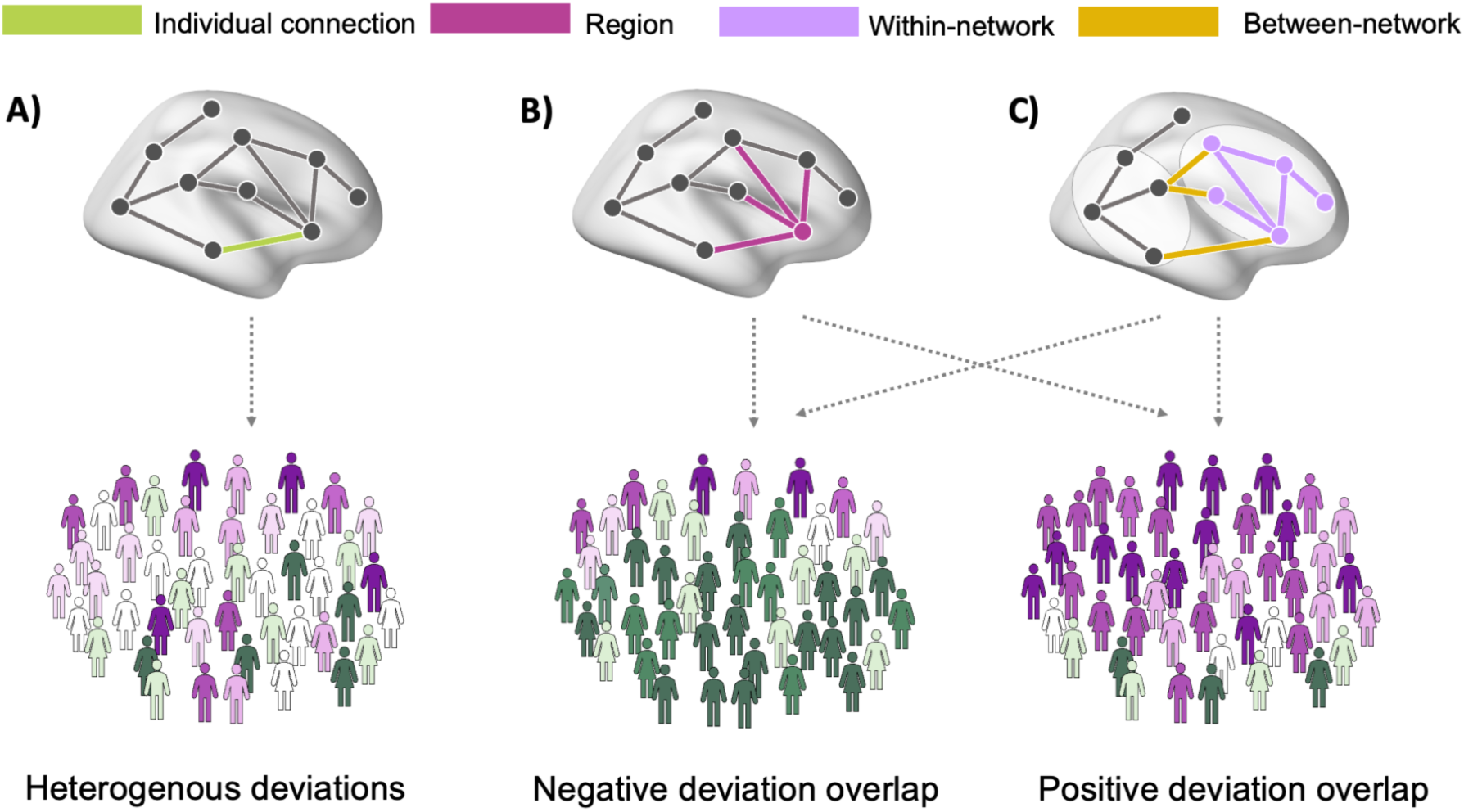
Multi-scale heterogeneity of functional connectivity deviations in autism. A) Connection-level analysis involves quantifying the extent to which each person deviates from model expectations at each inter-regional FC estimate. We expect a high degree of heterogeneity in which specific connections are deviant in each individual. B) Despite heterogeneity at the level of inter-regional connections, deviations may nonetheless converge on FC estimates attached to specific brain regions. We expect a higher level of inter-individual consistency at this regional level. C) FC deviations may also occur each within specific canonical networks, or between specific pairs of networks. Bottom panels show the greater prevalence of either positive (purple) or negative (green) deviations from model expectations within the autism sample.

## METHODS AND MATERIALS

### Participants

We pooled scans from three large datasets: the EU-AIMS Longitudinal European Autism Project (LEAP) (Loth et al., 2017) (https://www.eu-aims.eu/ and https://www.aims-2-trials.eu/), and the Autism Brain Imaging Data Exchanges I and II, or ABIDE-I and ABIDE-II (Adriana Di Martino et al., 2017; A. Di Martino et al., 2014) (http://fcon_1000.projects.nitrc.org/indi/abide/), details can be found in Supplement Section 1. Quality control and exclusion criteria are detailed in the Supplement Section 2 and follow previous work (Ilioska et al., 2023).

The final sample included 796 autistic individuals (141 females; age-range: 5-58) and 1028 neuro-typical (NT) individuals (256 females; age-range: 5-56) recruited across 32 different sites. Table 1 contains detailed information on the clinical and demographic characteristics of the participants included in the study.

**Table 1.** Demographic and clinical information. ADOS, Autism Diagnostic Observation Schedule; ADI-R, Autism Diagnostic Interview-Revised; autism, autism spectrum disorder; NT, neurotypicals; FD, framewise displacement; RRB, Restrictive Interests and Repetitive Behavior; SRS-2, Social Responsiveness Scale Second Edition; AQ, Autism Spectrum Quotient "-", not applicable; FD, framewise displacement; SD, standard deviation

|  | EU-AIMS LEAP |  |  | ABIDE 1 |  |  | ABIDE 2 |  |  | ALL |  |  |
| --- | --- | --- | --- | --- | --- | --- | --- | --- | --- | --- | --- | --- |
|  | Autism | NT | t-value,<br>p-value | Autism | NT | t-value,<br>p-value | Autism | NT | t-value,<br>p-value | Autism | NT | t-value,<br>p-value |
| n | 222 | 200 | .. | 369 | 481 | .. | 205 | 347 | .. | 796 | 1028 | .. |
| Male/female<sup>a</sup> | 158/64 | 131/69 | 1.31,<br>0.251 | 321/48 | 390/91 | **4.9,**<br>**0.026** | 176/29 | 251/96 | **12.68,**<br>**0.0003** | 655/141 | 772/256 | **13.2,**<br>**0.0002** |
| Age<br>(mean ± SD) | 18.1 ±5.7 | 18.3<br>±5.8 | -0.4,<br>0.7 | 17.0<br>±7.8 | 17.1<br>±7.3 | -0.3, 0.8 | 14.0<br>±7.0 | 13.0<br>±5.74<br>23 | 1.931,<br>0.0539 | 16.2<br>±7.0 | 15.95<br>±6.9 | 1.7, 0.1 |
| Full-scale IQ<br>(mean ± SD,<br>n) | 105.5<br>±15.2,<br>221 | 108.5<br>±12.6,<br>199 | **-2.2,**<br>**0.028** | 106.4<br>±16.3,<br>361 | 111.4<br>±12.4,<br>464 | **-5.0,**<br>**<0.0001** | 106.1<br>±15.7,<br>195 | 115.4<br>±12.7<br>, 331 | **-7.4,**<br>**<0.0001** | 106.1<br>±15.8,<br>777 | 112.2<br>±12.8<br>, 994 | **-8.97,**<br>**<0.0001** |
| Head motion<br>meanFD<br>(mean ± SD) | 0.076<br>±0.043 | 0.069<br>±0.035 | 1.933,<br>0.054 | 0.081<br>±0.034 | 0.071<br>±0.031 | **4.41,**<br>**<0.0001** | 0.077<br>±0.035 | 0.075<br>±0.033 | 0.9,<br>0.365 | 0.079<br>±0.037 | 0.072<br>±0.033 | **4.18,**<br>**<0.0001** |
| Handedness<br>(right/left/am<br>bidextrous,<br>n) | 158/26/6,<br>190 | 144/1<br>5/4,<br>163 | .. | 207/30/<br>6, 243 | 300/25/<br>6, 331 | .. | 158/17/<br>19, 194 | 310/1<br>6/14/<br>340 | .. | 523/73/<br>31, 627 | 754/5<br>6/24,<br>834 | .. |
| Current<br>medication<br>use | 84 | 12 | .. | 85 | 2 | .. | 38 | 7 | .. | 207 | 21 | .. |
| ADOS social<br>affect (mean<br>± SD, n) | 5.7 ±2.5,<br>217 | .. | .. | 8.8<br>±4.2,<br>189 | .. | .. | 9.3<br>±3.7,<br>105 | .. | .. | 7.6<br>±3.9,<br>509 | .. | .. |
| ADOS RRB<br>(mean ± SD,<br>n) | 4.5<br>±2.6,<br>217 | .. | .. | 2.7<br>±1.8,<br>187 | .. | .. | 3.0<br>±1.8,<br>105 | .. | .. | 3.5<br>±2.3,<br>511 | .. | .. |
| ADI social<br>(mean ± SD,<br>n) | 15.7<br>±6.6, 212 | .. | .. | 19.8<br>±5.3,<br>255 | .. | .. | 18.4<br>±5.3,<br>117 | .. | .. | 18.0<br>±6.1,<br>584 | .. | .. |
| ADI<br>communication (mean ±<br>SD, n) | 12.9<br>±5.6, 212 | .. | .. | 15.9<br>±4.5,<br>256 | .. | .. | 14.8<br>±4.5,<br>116 | .. | .. | 14.6<br>±5.1,<br>584 | - | .. |
| ADI RRB<br>(mean ± SD,<br>n) | 3.9 ±2.5,<br>212 | .. | .. | 6.0<br>±2.5,<br>256 | .. | .. | 5.3<br>±2.3,<br>117 | .. | .. | 5.1<br>±2.6,<br>585 | .. | .. |
| SSP Hypo.<br>(mean ± SD,<br>n) | 31.4<br>±6.9,<br>140 | 37.6<br>±3.6,<br>82 | **-7.4,**<br>**<0.0001** | .. | .. | .. | 5.3<br>±2.3,<br>117 | .. | .. | 31.4<br>±6.9,<br>140 | 37.6<br>±3.6,<br>82 | **-7.4,**<br>**<0.0001** |
| SSP Hyper.<br>(mean ± SD,<br>n) | 54.3<br>±10.4,<br>137 | 67.4<br>±3.9,<br>83 | **-10.9,**<br>**<0.0001** | 6.0<br>±2.5,<br>256 | .. | .. | 5.3<br>±2.3,<br>117 | .. | .. | 54.3<br>±10.4,<br>137 | 67.4<br>±3.9,<br>83 | **-10.9,**<br>**<0.0001** |
| SRS t-score<br>(mean ± SD,<br>n) | 85.2<br>±30.2,<br>178 | 19.9<br>±14.3,<br>93 | **19.74,**<br>**<0.0001** | 91.0<br>±30.3,<br>160 | 21.9<br>±16.8,<br>165 | **25.5,**<br>**<0.0001** | 86.6<br>±30.20,<br>170 | 20.0<br>±15.0<br>, 277 | **31.0,**<br>**<0.0001** | 87.5<br>±30.3,<br>508 | 20.6<br>±15.4<br>, 535 | **45.33,**<br>**<0.0001** |
| **AQ**<br>(mean ± SD,<br>n) | 89.7<br>$\pm 20.6$,<br>181 | 43.8<br>$\pm 16$,<br>158 | **22.6**,<br>**<0.0001** | 29.2<br>$\pm 9.0$,<br>15 | 12.8<br>$\pm 6.2$,<br>15 | **5.7**,<br>**<0.0001** | 32.4<br>$\pm 9.38$,<br>20 | 17.25<br>$\pm 4.2$,<br>20 | **31.0**,<br>**<0.0001** | 80.1<br>$\pm 28.9$,<br>216 | 38.6<br>$\pm 18.2$,<br>, 193 | **17.11**,<br>**<0.0001** |

### Clinical diagnosis

Autistic individuals in the EU-AIMS LEAP were clinically diagnosed with autism according to DSM-IV, DSM-5, or ICD-10 criteria. Most of these individuals also meet criteria for autism on the Autism Diagnostic Interview-Revised (ADI-R) (Rutter, Le Couteur, & Lord, 2003) and/or the Autism Diagnostic Observation Schedule 2 (ADOS 2) (C Lord et al., 2012) tests. For more details on the clinical diagnosis procedure in LEAP, see (Charman et al., 2017). Each research site in the ABIDE study used their own procedures for diagnosing autism, although the majority of the sites used the Autism Diagnostic Observation Schedule (C. Lord et al., 2000) and/or the Autism Diagnostic Interview-Revised (Rutter et al., 2003). Neuro-typical controls are individuals who do not have a psychiatric diagnosis.

### MRI acquisition and Pre-processing

Resting-state fMRI and structural scans were obtained using 3T MRI scanners at 32 scanning sites. For more details on the MRI acquisition and denoising procedures see Supplement Section 3 and 4.

### Mapping functional connectivity

To map inter-regional FC, we used the well-validated Schaefer parcellation (Schaefer et al., 2018) to obtain 400 functional cortical regions of interest (ROIs) for each participant. We also included 15 subcortical ROIs from the Harvard-Oxford subcortical atlas (Craddock, James, Holtzheimer, Hu, & Mayberg, 2012). A total of 24 regions were excluded due to low coverage (<70% coverage in >5% of the participants) leaving 390 regions remaining for analysis (Figure S1). Within the Schaefer parcellation, each brain region is assigned to one of seven canonical functional networks, as identified in (Yeo et al., 2011). Subcortical regions were labelled as thalamus, striatum and medial temporal lobe.

We used Pearson correlations to calculate the relationship between each pair of regional average time series. We then normalized the values within each participant’s FC matrix using Gaussian-gamma mixture modeling (Llera, Vidaurre, Pruim, & Beckmann, 2016). This step functions as a soft thresholding method, enhancing the differentiation between meaningful correlations and statistical noise (Supplement section 5). We used ComBat (Johnson, Li, & Rabinovic, 2007) to regress the scan-site-specific effects from the FC data (for a confirmation of the success of this method, see Supplement Section 6).

### Normative modeling

We used gaussian process regression, as described by (Marquand et al., 2016), to fit separate normative models to each of 75,855 FC estimates between each pair of 390 brain regions, using age, sex, and mean framewise displacement. This method allows for the calculation of non-linear models that can adaptively fit complex datasets. Deviations from the normative model were calculated by subtracting the predicted from the observed FC and dividing by the estimated variance at each of the 75,855 connections between 390 parcellated brain regions. The normative model was trained on the group of neurotypical individuals to establish a normative range for FC and was then tested on the FC maps of the autistic individual’s group to quantify the extent of FC deviations from the normative baseline. We quantified the extent of FC deviations in the control group using 10-fold cross-validation. Calibration and evaluation metrics of the model are presented in Supplement Section 7.

### Connection-level analysis

We applied a threshold of |Z|>2.3 (corresponding to probability p=0.01) to the individual FC deviation maps to identify extreme positive and negative deviations from the normative model for each individual. To assess differences between the total number of extreme deviations in autistic individuals and neurotypical controls at any given connection, we used a Wilcoxon rank-sum test (Wilcoxon, 1945). This test was performed for both positive and negative deviations from the normative model.

We further sought to investigate whether the two groups have different ratios of positive to negative deviations. We calculated the within-participant ratio of positive to negative deviations across all connections and used an independent, two sample t-test to compare the ratios between the two groups of participants (autistic and neurotypical). This allowed us to assess whether the FC in autistic individuals is biased towards one polarity of extreme deviations compared to neurotypical individuals.

Next, we compared the degree of inter-individual heterogeneity by quantifying the difference in overlap in connection-level FC deviations between autistic individuals and neurotypical individuals. We calculated the percentage of people within each group showing extreme deviations at each connection (Figure 2A). To assess the significance of the group difference, we shuffled group labels 10,000 times and re-estimated the percentage overlap at each connection under the null hypothesis of random group assignment. We compared the original difference to the null, to determine the *p*-value at each connection and the Benjamini-Hochberg false discovery rate (FDR_BH_) method to correct for multiple comparisons (Benjamini & Hochberg, 1995).

**Figure 2.**
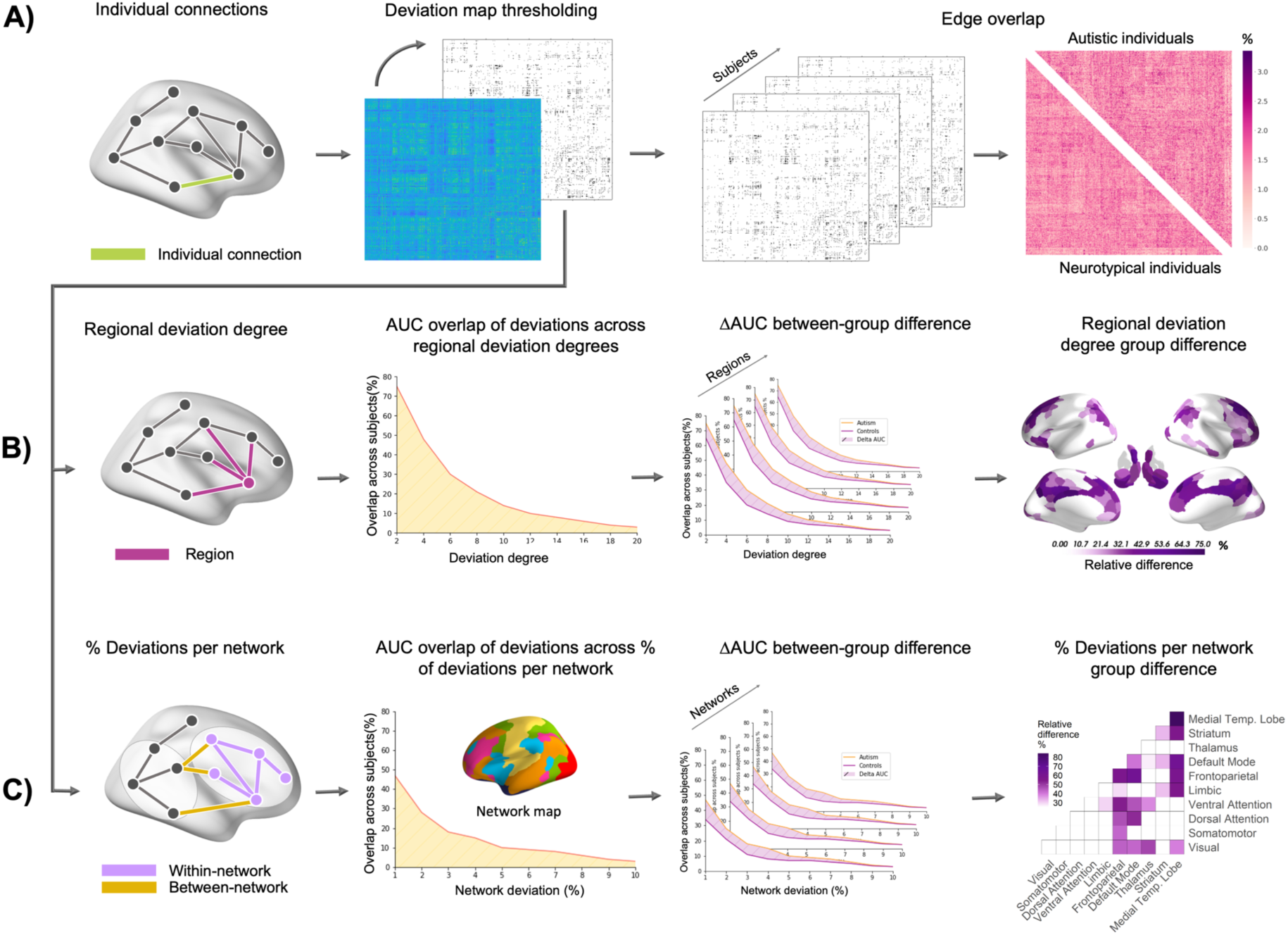
Workflow for quantifying deviation overlap across participants at the levels of connections, regions, and networks. A) Using the normative modeling pipeline, we generated a deviation map for each participant, comprising deviation estimates for 75,855 connectivity estimates between each pair of 390 regions. A z-score threshold of 2.3 (p = 0.01) was used to identify extreme deviations. The thresholded matrices were subsequently used to calculate the overlap in extreme deviations at each connection across participants. B) In the region level analyses, we quantified the regional deviation degree as the total number of extreme deviant FC estimates attached to each region. We then applied a threshold to the resulting vector using values ranging from 1 to 20 deviant connections and calculated the number of individuals with a regional deviation degree equal to or exceeding each threshold in each region. We computed the area under the curve (AUC) across these thresholds for each region in each group. We then calculated the difference ΔAUC between groups for each region and performed inference with a permutation test. C) A similar approach was used at the network level. We calculated the percentage of extreme FC deviations falling either within or between each pair of 10 canonical networks. We thresholded the resulting matrix using thresholds between 1% and 10%, and used the AUC coupled with permutation tests to compare groups.

### Region-level analysis

We computed the regional deviation degree of every region in each individual by summing the rows of their thresholded FC deviation matrix (Figure 2B). Regional deviation degree quantifies the number of extremely deviant connections attached to a region. We then thresholded the resulting regional deviation degree vectors of each participant at values ranging between 1 to 20, with a step of 1, and computed*O_R,G_*(τ) for region *R*, at each threshold, τ, as the number of participants within group *G* with a regional deviation degree higher than τ. The upper bound of 20 was chosen because at this threshold the number of subjects overlapping approaches zero (Figure 3B). To avoid relying on a specific threshold, we calculated the area under the curve *AUC_R,G_* of the overlap across thresholds for every region, separately in each group. *AUC_R,G_* can be expressed as:

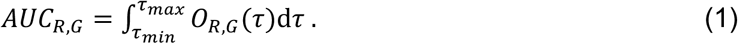

**Figure 3.**
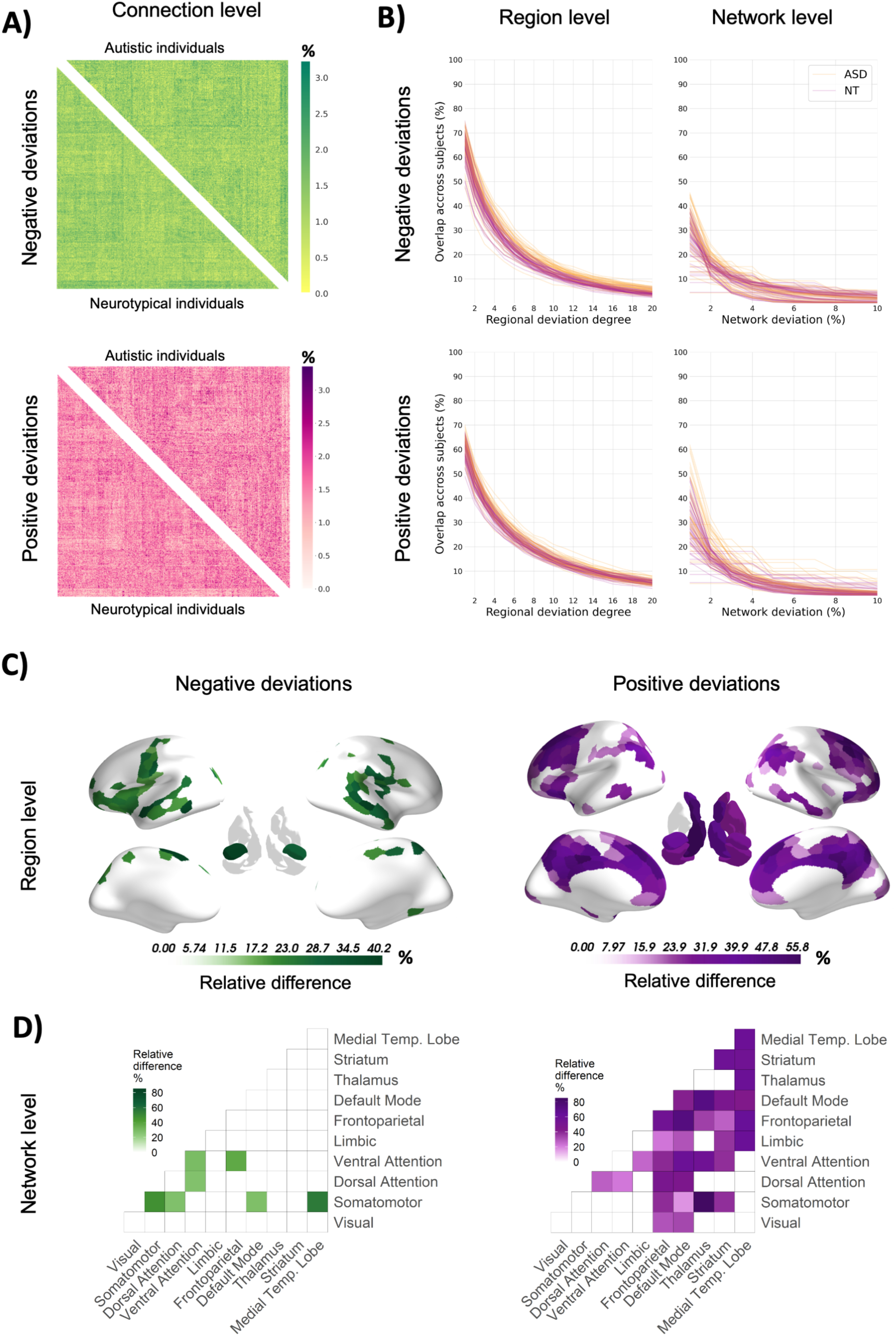
Group differences in deviation overlap at connection, region, and network levels. A) Connection level heatmap showing the percent overlap of FC deviations across participants (positively deviating – pink; negatively deviating – green). Positive deviations are connections with larger value than expected based on the normative model whereas negative deviations are connections that are lower than predicted by the normative model. The upper triangle shows the overlap across autistic individuals, whereas the lower triangle shows the overlap across neurotypical controls. B) Line plots showing the overlap of participants at the region and network levels. The first column shows the overlap for positive and negative regional deviation degree across 20 thresholds. Each line represents a region in the brain. Yellow color indicates overlap across autistic individuals, whereas red indicates overlap across neurotypical controls. The second column shows the percentage overlap of negative and positive network deviations. Here, each line represents a within or between network overlap across participants, where yellow indicates overlap across the group of autistic individuals and red indicates the group of controls. C), D) Regions and networks showing significantly greater overlap in autistic individuals represented as relative differences. Green color indicates overlap of negative deviations; purple color indicates overlap of positive deviations. Between-group differences are presented as the percentage difference in the overlap between the groups.

We subsequently calculated the difference Δ*AUC_R_* between the two groups and assessed the significance of the difference against a null distribution of Δ*AUC_R_*S derived from 10,000 permutations with randomly shuffled diagnostic labels. Δ*AUC_R_* is expressed as:

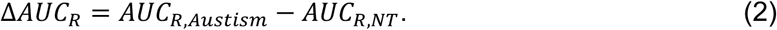

The p-values were FDR_BH_ corrected. This approach allowed us to compare group differences in deviation overlap at each region without having to rely on any single threshold value.

### Network-level analysis

The same AUC-based procedure was used to evaluate group differences in the overlap of network-level extreme deviations, thus quantifying the network-level deviation degree. At this level, we calculated the percentage of extreme deviations falling within or between canonical functional networks (Figure 2C). We then evaluated overlaps across thresholds ranging between 1% and 10% with steps of 1% (10% was chosen as the upper bound because the overlap of subjects at this threshold approaches zero for most networks (Figure 3B) and calculated the difference between the groups Δ*AUC_+_*, which in an expanded form can be expressed as:

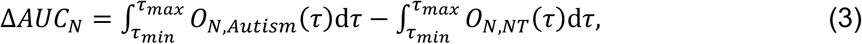

where *O_N,Autism_*(τ) is the overlap for threshold τ for the network *N* in the group of autistic individuals, and *O_N_*_,*NT*_ (τ) is the overlap for the network *N* in the group of neurotypical controls, for threshold τ. Inference was performed with 10,000 permutations and FDR_BH_-corrected *p*-values.

### Predicting clinical and cognitive variables from FC deviations

We used support vector regression (SVR) to develop multivariate predictive models of clinical and cognitive measures using FC deviation scores at the connection, region, and network levels. We used a diverse set of behavioral variables including the ADOS social affect, ADOS restricted and repetitive behavior (RRB), ADI RRB (C Lord et al., 2012), ADI communication, ADI social interaction (Rutter et al., 2003), Social Responsiveness Scale-2 (SRS) (Constantino & Gruber, 2012), full scale IQ from the Wechsler Abbreviated Scales of Intelligence—Second Edition (Wechsler, 1999) and the Short Sensory Profile scale (SSP) (McIntosh, Miller, Shyu, & Dunn, 1999) (Lefebvre et al., 2023) and Autism Quotient (AQ) (Baron-Cohen, Wheelwright, Skinner, Martin, & Clubley, 2001). For descriptive statistics of the variables see Table 1. For more information on the variables see Supplement Section 8.

Separate models were run for each variable and each FC resolution. For each model, we used a five-fold cross validation procedure where each fold split the data into a discovery (70% of the data in the fold) and validation dataset. We applied a principal component analysis (PCA) to the discovery data. To reduce dimensionality, we retained the principal components that explained 50% of the variance. We trained a linear SVR, epsilon insensitive model using unthresholded deviation values to assess the connection-level relationships, while network and regional values were calculated on data thresholded at τ=2.3, as in the overlap analysis. Five-fold grid-search cross validation was applied to find the best ε and C parameters according to the coefficient of determination. The held-out validation data were projected into the discovery data principal component space, to ensure that the model trained on the discovery set generalizes to unseen data. We next applied the linear SVR model on the PCA-projected validation data sets to predict the phenotypic scores in unseen data. The strength of the association in out-of-sample data is quantified by the correlation between the model’s predicted scores and the observed phenotypic scores. We report the median correlation across the five folds and the interquartile range of the five folds. The entire procedure was repeated 1,000 times with shuffled order of the subjects on one side, to assess the significance of the model’s predictions. This procedure was performed separately for each variable and each level of FC deviations. The resulting p-values were corrected using the false discovery rate (FDR) correction (Benjamini & Hochberg, 1995).

## RESULTS

### Heterogeneity at the level of single connections

The total number of extreme positive deviations in FC identified across individuals with ASD ranged between 0 and 5652 deviations, with a median of 741.5. The number of extreme negative deviations ranged between and 0 and 5119, with a median of 657. The total number for positive deviations in controls ranged between 35 and 3570 with a median of 760, and the number of negative deviations between 105 and 4745, with a median 673.5.

The Wilcoxon rank-sum test did not show significant group differences in the median number of extreme positive or negative deviations (p_positive_=0.086; p_negative_=0.4). The two groups also showed no significant differences in the ratio of positive-to-negative deviations (t=0.58; p=0.55).

The level of overlap observed at any individual connection was minimal and did not exceed 3.4% in either group (Figure 3A). Permutation testing did not uncover any significant group differences in connection-level overlap for either positive or negative deviations (all *p_FDR_*>.05). These findings indicate that autism is associated with extreme heterogeneity in the specific pair-wise connections showing atypical FC.

### Heterogeneity at the level of individual brain regions

We observed higher overlap across participants for both groups at the region level, for both positive and negative extreme deviations. The level overlap ranged between 41.9% and 77.8% across brain regions for the lowest threshold of t=1. At the maximum regional deviation degree threshold of t=20, the overlap decreased to between 1.8% and 12.8% (Figure 3B). Statistical comparison of regional deviation degree overlap across thresholds in each region revealed significantly higher overlap in autistic individuals compared to neurotypical for negative deviations in sensory-motor, anterior insula, prefrontal, temporal pole, visual, and medial temporal regions (Figure 3C). Autistic individuals showed higher overlap for positive deviations in medial prefrontal, superior frontal, cingulate, inferior parietal lobules, and subcortical areas (Figure 3C). Controls did not show significantly greater overlap in any region. For results obtained using a more liberal threshold for defining extreme deviations of t=1.7 and a more conservative threshold of t=3.1, see Supplement Section 9.

### Heterogeneity at the level of canonical functional networks

Overlap at the network level ranged between 4.4 and 61.9% at the lowest deviation degree threshold of t=1. At the highest threshold, the overlap ranged between 0 and 5.4% (Figure 3B). Autistic individuals showed significantly greater overlap for negative deviations within the visual, ventral attention, limbic, and default mode networks, as well as for connections linking the somatomotor network to other systems; namely, the visual dorsal attention, and ventral attention networks. Overlap in autism was also higher for connections linking medial temporal regions with the somatomotor, ventral attention, limbic, and default mode networks.

Greater overlap in autism for positive deviations was primarily concentrated in connections linking the default mode network and cognitive control networks with the rest of the brain, as well as subcortical-subcortical and subcortical-cortical connections (Figure 3C).

Controls did not show significantly greater overlap at the network level. See Supplement Section 9 for results with thresholds 1.7 and 3.1.

### Sensitivity analyses

Sensitivity analyses indicated that our findings were robust to the effects of participant sex, ADHD comorbidity, and psychotropic medication (see Supplement Section 10 and Table S4).

### Prediction of clinical and cognitive measures

Figure 4 shows the median correlations between true and predicted values for the modelled variables across the three levels. Single connections were significantly predictive of full-scale IQ (r_median_=0.16; p_FDR_<0.001) and SRS (r_median_ =0.29; p_FDR_<0.001). At the regional level, deviations showed a significant prediction of full-scale IQ (r_median_=0.1; p_FDR_=0.01) and SRS (r_median_ =0.1; p_FDR_= 0.02). The network-level model significantly predicted ADI social (r_median_ =0.17; p_FDR_<0.001), AQ (r_median_ =0.17; p_FDR_<0.02) and SRS (r_median_ =0.18; p_FDR_<0.001). These results suggest that different levels of network organization are related to distinct clinical features of autism.

**Figure 4.**
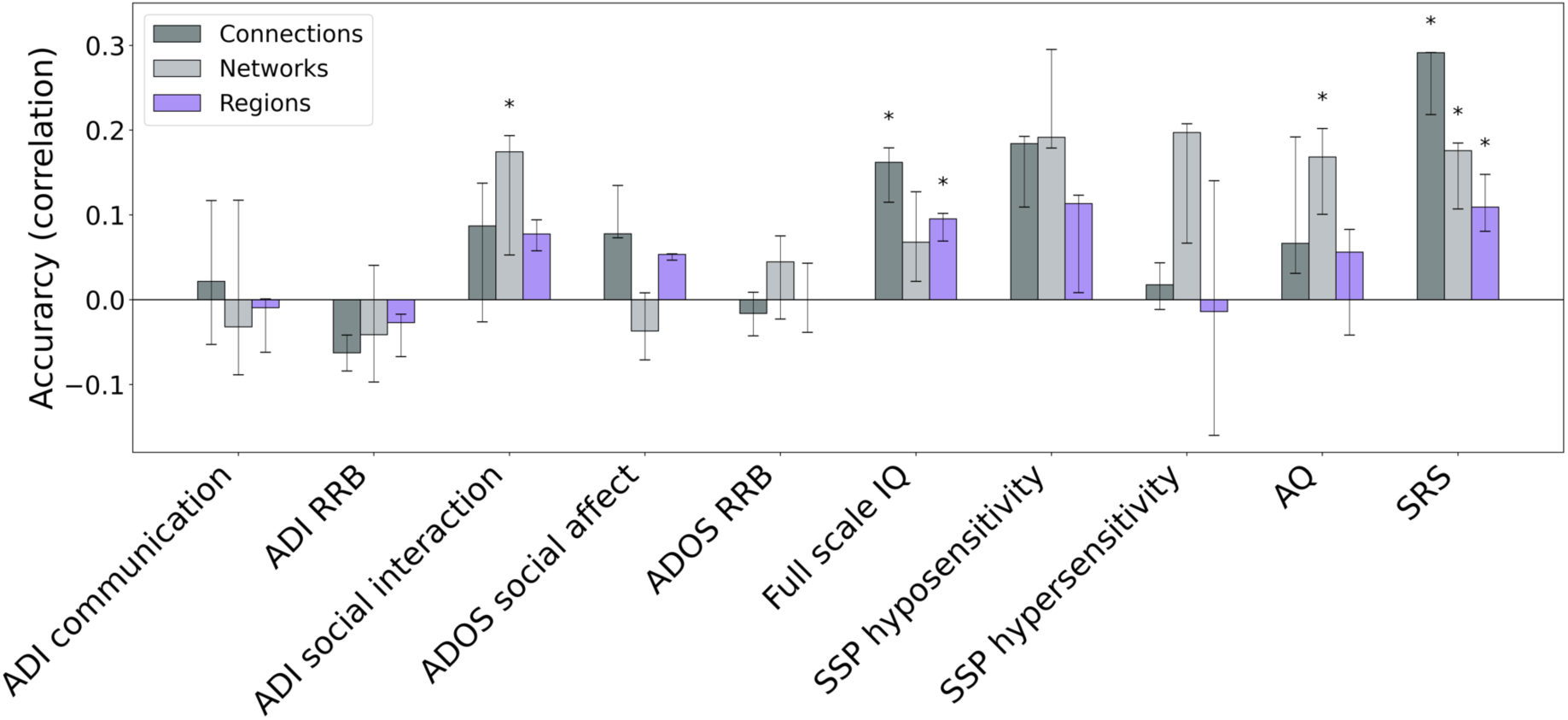
Support vector regression correlations of true and predicted values across levels of FC deviations and variables. Median accuracy across 5 folds of the analysis. The error bars represent the variability of the result across folds (inter-quartile range). We used scales from Autism Diagnostic Interview (ADI), Autism Diagnostic Observation Schedule (ADOS), Wechsler Abbreviated Scales of Intelligence the Short Sensory Profile (SSP), Autism spectrum Quotient (AQ) and the Social Responsiveness Scale (SRS).

## DISCUSSION

We used normative modelling to move beyond group mean comparisons and characterize the inter-individual heterogeneity of FC in autism at the connection, region, and network levels. We hypothesized that heterogeneous deviations at the most granular level of single connections would converge on common brain regions and networks in autism. In accordance with our hypothesis, we found evidence of substantial heterogeneity at the connection level, such that no more than 3.4% of autistic or neurotypical individuals showed an extreme deviation at the same connection. Overlap at the region and network levels was much higher, with autistic individuals showing greater overlap of negative deviations in sensory-motor areas, anterior insula, prefrontal, temporal pole, visual, and medial temporal regions, as well as the somatomotor, and ventral attention networks. They also showed greater positive deviation overlap in medial prefrontal, superior frontal, cingulate cortex, inferior parietal lobules, and subcortical areas, as well as frontoparietal and default mode networks. Moreover, FC deviations predicted distinct behaviors at each level. Together, extreme connection-level heterogeneity and higher regional and network-level overlap represent plausible neural substrates for phenotypic differences and similarities between autistic individuals, respectively.

### Connection-level deviations are highly heterogeneous

The high level of connection-level heterogeneity suggests that classical group mean comparisons conducted at this granular scale are unlikely to be representative of autistic individuals. Studies examining grey matter volume and cortical thickness have not found significant overlap at the most detailed levels of analysis (Segal et al., 2022 ; Zabihi et al., 2019). This suggests that there is high heterogeneity when comparing results across different brain imaging modalities at their most fine-grained levels. This result may explain the inconsistency of FC findings reported in the literature, with reports of increased FC, decreased FC, or both, being common, often within the same systems (J. V. Hull et al., 2016). We also found no group differences in total connection-level deviation degree or the ratio of positive to negative extreme deviations indicating that autistic individuals show no evidence for either predominantly hypo- or hyperconnectivity, consistent with our earlier work indicating that autistic individuals exhibit a complex array of both hypo and hyperconnectivity (Ilioska et al., 2023).

### Heterogeneous FC deviations converge on common regions and networks

In contrast to the extreme heterogeneity at the level of inter-regional connections, we observed considerably higher overlap at the level of brain regions and canonical functional networks (Figure 3, C&D). This result aligns with recent work on grey matter volume indicating that while deviations of regional grey matter volume are located in highly heterogeneous areas, these deviations are often coupled to common circuits and networks across autistic individuals(Segal et al., 2022).

Negative FC deviations (i.e., atypically reduced FC) generally showed greater regional and network-level overlap in autistic individuals in somatomotor, frontal, and temporal regions. At the network level, autistic individuals also showed greater deviation overlap for connections linking the somatomotor system with the dorsal and ventral attentional systems, the medial temporal lobe, and the DMN. Atypical sensory processing and motor coordination are well documented in the autism literature (Fournier, Hass, Naik, Lodha, & Cauraugh, 2010; Marco, Hinkley, Hill, & Nagarajan, 2011). This result aligns with past work (Ilioska et al., 2023; Mostofsky et al., 2009; Nebel, Eloyan, Barber, & Mostofsky, 2014 ; Nebel, Joel, et al., 2014) showing reduced FC within sensory-motor areas and between sensory-motor areas and attentional systems. Weaker connectivity between these networks was associated with social difficulties and restricted and repetitive behaviors in previous work (Ilioska et al., 2023).

Autistic individuals showed greater overlap for positive deviations (i.e., higher connectivity than expected) within the default mode network and between the default mode and other systems. Previous literature has comprehensively documented the connectivity patterns of the default mode network (Assaf et al., 2010; Monk et al., 2009; Padmanabhan, Lynch, Schaer, & Menon, 2017; Supekar et al., 2013). Our observation of increased overlap of positive FC deviations in the DMN highlights the complex role of the DMN in the neural underpinnings of the behavioral phenotype in autism. The DMN is closely associated with self-referential thought and is most active during periods of rest and introspection (Raichle, 2015). Increased connectivity of the DMN with other brain regions may contribute to a more inward-focused cognitive experience and a reduced inclination to engage with the external environment (Buckner, Andrews-Hanna, & Schacter, 2008; Raichle, 2015).This heightened connectivity can potentially explain tendencies towards introspection and diminished social interaction and sensory processing aligning with the reduced connectivity of sensory systems (Qin & Northoff, 2011; Whitfield-Gabrieli & Ford, 2012).

Positive deviations also exhibited greater overlap for frontoparietal network FC. This result aligns with studies reporting hyperconnectivity in the frontoparietal or cognitive control network in autistic individuals (Supekar et al., 2013; Uddin et al., 2015). It suggests that these deviations could be associated with the behavioral inflexibility often seen in autistic individuals, as this network plays a key role in cognitive control and task switching (Kenworthy, Yerys, Anthony, & Wallace, 2008; Supekar et al., 2013; Uddin et al., 2015). We found that positive deviations also occurred more frequently between subcortical and cortical brain areas in autistic individuals, in line with past research showing increased connectivity in these systems (Cerliani et al., 2015; A. Di Martino et al., 2014; Ilioska et al., 2023).

Collectively, these findings suggest that a core neural phenotype of autism that is shared across different autistic individuals involves reduced FC of somatomotor areas and increased FC of higher-order trans-modal areas on the level of regions and networks, consistent with reports of altered hierarchical function in autistic individuals (Hong et al., 2019) and degree centrality findings (Holiga et al., 2019; Ilioska et al., 2023). However, we emphasize that this phenotype, fully or partially, may only be common across a subgroup of autistic individuals. It will be important to understand common and divergent characteristics of autistic individuals with those who do not share this phenotype. Such an analysis will require very large samples to appropriately parse the heterogeneity of deviations with the relevant neural systems.

### Clinical and behavioral predictions

Despite the high degree of heterogeneity at the connection level, FC deviations significantly predicted intellectual ability and social functioning. Regional deviation degree also significantly predicted intellectual ability, highlighting that intellectual ability is captured at both the fine-grain level of single connections and the coarser level in which FC deviations converge on specific brain regions.

Scores from the SRS were significantly predicted by deviation patterns at all levels, while the ADI social functioning scale was predicted by network-level FC deviations. Both scales capture different dimensions of social functioning. The SRS reflects broader social responsiveness, including social awareness, social cognition, social motivation, and communication (Constantino & Gruber, 2012). In contrast, the ADI social functioning scale assesses specific aspects of social interaction related to autism, such as reciprocal social interaction and peer relationships (Rutter et al., 2003). Both scales have been previously linked to FC. Recent work found that inter-regional and voxel-wise FC within brain circuits responsible for social cognition negatively correlated with SRS scale scores (Wang, 2023).

Additionally, the SRS was associated with complex patterns of both increased and decreased connectivity at the network level (Oldehinkel et al., 2019). ADI social interaction was both positively and negatively related to functional connectivity between various brain regions in (Bathelt, Geurts, & Borsboom, 2022). Additionally, in a recent work, sociability was positively linked to somatosensory connectivity, which aligns with the overlap pattern of negative deviations in autism and the association of the region and network level deviations with social difficulties (Rovný et al., 2024). Together these relationships suggest that FC deviations reflect social functioning at all levels.

Similarly to the ADI social functioning scale, the AQ scale was predicted solely by deviations at the level of networks. The AQ is a self- or parent-administered questionnaire designed to measure “broader phenotype” of autistic traits, capturing social skills, communication, imagination, attention to detail and task-switching ability. The instrument is not designed for diagnostic purposes. Rather it is intended to indicate whether a person should pursue further diagnostic assessment (Baron-Cohen et al., 2001). Variability in general autistic traits may therefore result from the global macro-organization of FC.

### Limitations and conclusions

Our analysis approach relies on the choice of a threshold for (a) defining extreme deviation at the connection level; and (b) computing overlap at the region and network-levels. We showed that our general conclusions are robust to the specific choices made for these thresholds, but the threshold chosen will necessarily influence the exact levels of overlap observed across individuals.

Due to poor scan coverage in a large portion of the participants, our analysis did not include the cerebellum, which is thought to play an important role in autism pathophysiology (D’Mello & Stoodley, 2015; Khan et al., 2015; Oldehinkel et al., 2019). Our sample also had an imbalanced sex ratio between the autism and control groups, which aligns with the higher prevalence of autism in males compared to females. Future research should prioritize including more autistic females to better understand the unique characteristics and needs of this group.

In summary, our results highlight the importance of adopting a multiscale approach to characterizing the heterogeneity of neural phenotypes in autism. Extreme heterogeneity at the level of specific connections offers a plausible neural substrate for clinical differences between autistic individuals and may explain the inconsistent array of FC findings reported in the literature thus far. Higher overlap at the regional and network levels represents a plausible substrate for clinical similarities between autistic individuals. Our findings further suggest that reduced FC of somatomotor systems and increased FC of transmodal association networks, potentially reflecting imbalanced signaling along the sensorimotor-association axis of the brain, may represent a core phenotype that is shared across a large fraction of autistic individuals. FC deviations at distinct levels predict different clinical phenotypes, emphasizing the importance of considering multiple levels when characterizing brain-behavior relationships.

## Data Availability

Data from EU-AIMS LEAP is available upon application and approval to the EU-AIMS LEAP committee (https://www.eu-aims.eu/). Data from the ABIDE initiative is publicly available (https://fcon_1000.projects.nitrc.org/indi/abide/)

