## Supplemental Material for "Multiscale heterogeneity of functional connectivity in autism"

**Section 1. Data**
We used data from three consortia: the EU-AIMS Longitudinal European Autism Project (LEAP), accessible at (https://www.eu-aims.eu/ and https://www.aims-2-trials.eu/), and the Autism Brain Imaging Data Exchange (ABIDE) 1 and ABIDE 2, available at (http://fcon_1000.projects.nitrc.org/indi/abide/).

The LEAP consortium focuses on the identification and classification of autism biomarkers across multiple research centers (Loth et al., 2017). A total of 437 autistic and 300 neurotypical individuals were recruited across five sites. Male and female participants, aged between 6 and 30 years, underwent standardized MRI protocols and comprehensive clinical, cognitive, and genetic assessments. For a more in-depth understanding of the LEAP study, refer to (Charman et al., 2017; Loth et al., 2017).

ABIDE is an open access initiative, providing access to collected rs-fMRI and structural MRI data from autistic participants and matched controls (Adriana Di Martino et al., 2017; A. Di Martino et al., 2014). Individual studies were initially conducted at various sites before being combined into a single dataset. ABIDE 2 expands upon the ABIDE 1 initiative, and collectively, these efforts offer 2,226 unique datasets from 32 sites, including data from 1,060 autistic individuals and 1,166 controls aged between 5 and 64.

**Section 2. Exclusion Criteria**
Initially, the LEAP dataset contained 617 participants with available demographic and rs-fMRI data. Subjects with low-quality resting-state fMRI scans due to preprocessing failure (N=2) and those with incomplete scans (N=8) were excluded. To account for head motion contamination during the resting-state fMRI scan, participants were excluded if they met at least one of the following criteria: a mean framewise-displacement (mFD) > 0.25mm, more than 20% of FDs above 0.2mm, or any FDs larger than 5mm (N=123). These criteria are consistent with the "stringent" exclusion guidelines reported in previous research (Parkes, Fulcher, Yucel, & Fornito, 2018; T. D. Satterthwaite et al., 2013).

We next excluded parcellated ROIs with over 30% signal dropout in more than 5% of the entire subject sample (Figure S1) and subjects who had poor coverage (<70%) in more than 10% of the remaining regions. In the LEAP cohort, this procedure did not result in any exclusions (N=0).

Carpet plots for each participant were then visually inspected (Aquino, Fulcher, Parkes, Sabaroedin, & Fornito, 2020; Power, 2017). Displaying the complete fMRI time series in matrix form with a normalized color scale representing signal fluctuations at each time point in each voxel, these plots proved effective in identifying significant imaging artifacts and signal contributions from brain-wide signal changes. An additional 17 participants with noticeable artifacts were excluded based on these plots. All participants with an IQ < 70 (N=51) and those with structural brain abnormalities (N=4) were also excluded, resulting in a final total of 422 subjects for analysis.

In the ABIDE 1 cohort, demographic and resting-state fMRI data were available for 1,110 participants. Participants with low-quality scans due to preprocessing failure (N=9) and those with incomplete scans (N=8) were excluded. To account for excessive head motion during the resting-state fMRI scan, we applied the same criteria as in the LEAP dataset resulting in N=173 exclusions. A total of 20 participants were excluded for poor signal/anatomical coverage. Participants with visible carpet-plot-artifacts (N=8) were removed. All participants with IQ < 70 (N=22) and those with structural brain abnormalities (N=4) were also excluded, resulting in a final count of 852 subjects included in the analysis.

For the ABIDE 2 cohort, data from sites with TR > 800ms were considered, and demographic and resting-state fMRI data were available for 902 participants. We excluded participants with incomplete scans (N=209) and excessive head motion (N=130). No participants were excluded based on low coverage (N=0). Following visual inspection of functional data carpet plots (Power, 2017), participants with visible artifacts (N=9) were excluded. All participants with IQ < 70 (N=2) were also excluded, resulting in a final total of 552 subjects for analysis.

**Section 3. MRI Acquisition and Pre-processing**

MRI data in the LEAP consortium were collected at five locations: Cambridge University, King's College London (KCL), Central Institute of Mental Health Mannheim, Radboud University Nijmegen Medical Centre, and University Medical Center Utrecht, using a uniform MRI protocol. A magnetization prepared rapid gradient-echo (MPRAGE) sequence, following the ADNI 2 / GO protocol (http://adni.loni.usc.edu), was used for structural scans, and a multi-echo planar imaging sequence was employed for resting-state fMRI scans (Kundu, Inati, Evans, Luh, & Bandettini, 2012). Table S1 provides further information on scanning acquisition parameters for the LEAP cohort.

For the ABIDE 1 cohort, data from sixteen sites were analyzed, including: California Institute of Technology (Caltech), Carnegie Melon University (CMU), Kennedy Krieger Institute (KKI), University of Leuven (Leuven), Ludwig Maximilian’s University Munich (MaxMun), NYU Langone Medical Center (NYU), Olin Institute of Living at Hartford Hospital (Olin), University of Pittsburgh School of Medicine (Pitt), Social Brain Lab BCN Neuroimaging Center, University Medical Center Groningen (SBL), San Diego State University (SDSU), Stanford University (Stanford), Trinity Centre for Health Sciences (Trinity), University of California Los Angeles (UCLA), University of Michigan (UM), University of Utah School of Medicine (USM), and Yale School of Medicine—Yale Child Study Center (Yale). Table S2 offers a detailed overview of acquisition parameters for each ABIDE 1 site.

Thirteen sites contributed data to the ABIDE 2 cohort analysis: Erasmus University Medical Center (EMC), Rotterdam, ETH Zürich (ETH), Georgetown University (GU), Indiana University (IU), Kennedy Krieger Institute (KKI), Katholieke Universiteit Leuven (KUL), NYU Langone Medical Center Sample 1 (NYU1), NYU Langone Medical Center Sample 2 (NYU2), San Diego State University (SDSU), Stanford University (Stanford), Trinity Centre for Health Sciences (TCD), University of California David (UCD), University of Utah School of Medicine (USM), and University of Miami (Miami). Table S3 provides further details on acquisition parameters for each ABIDE 2 sites.

All rs-fMRI images underwent preprocessing using the FMRIB Software Library (FSL; www.fmrib.ox.ac.uk/fsl) (Smith et al., 2004). The following pipeline was implemented: removal of the initial five volumes for signal equilibration, volume realignment to the median volume using MCFLIRT for primary head motion correction, grand mean scaling, and spatial smoothing employing a 6mm FWHM Gaussian kernel. ICA-AROMA was then used for secondary head motion-related artifact correction (Pruim, Mennes, van Rooij, et al., 2015). ICA-AROMA is capable of effectively removing motion-related artifacts while preserving neurobiological signals of interest (Pruim, Mennes, Buitelaar, & Beckmann, 2015), and has compares favorably to alternative methods for motion-related confound removal (Parkes et al., 2018). Mean signals from the CSF and white matter were regressed out as nuisance covariates, and a 0.01Hz temporal high-pass filter was applied. Participant functional images were co-registered to their respective anatomical images using boundary-based registration in FSL FLIRT (Jenkinson, Bannister, Brady, & Smith, 2002; Jenkinson & Smith, 2001). High-resolution structural images were registered to MNI152 standard space using a 12-parameter affine transformation and further refined with a non-linear registration via FSL FNIRT, employing a 10mm warp and 2mm resampling resolution (Smith et al., 2004). Finally, functional images were normalized to 2mm MNI152 standard space by applying the transformation of the functional image to T1 and T1 to MNI152. All subsequent analyses were conducted in MNI152 standard space.

We additionally excluded a further 57 participants from sites with very few (i.e., <10) female participants: the University of Leuven sample 2; Social Brain Lab, UMC Groningen; and the Erasmus Medical Center Rotterdam.

The final sample included 773 autistic individuals (138 females; age-range: 5-58) and 994 NT individuals (250 females; age-range: 5-56).

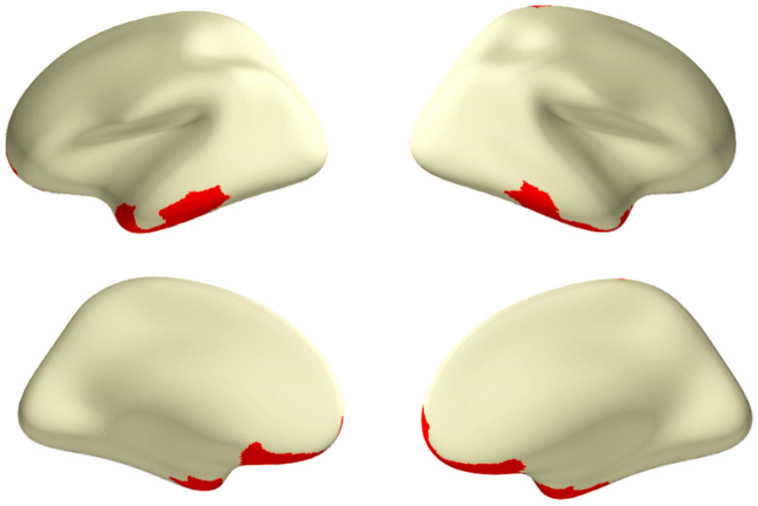

**Figure S1. ROIs excluded due to low coverage**
ROIs were omitted (red) when they exhibited low coverage (less than 70% of the voxels within the ROI) in over 5% of the participants.

**Section 4. Evaluation of Motion-Artifact Removal**

The quality control-functional connectivity (QC-FC) correlation denotes the association between the mean framewise displacement and functional connectivity for each edge. More specifically, it represents the correlation between observed FC values across participants and the participants' mean framewise displacement at each edge. This measure offers insights into the dependency of the FC value on head motion, reflecting the quality of the FC data. To estimate this relationship post-denoising, we computed the QC-FC correlations (Ciric et al., 2017; Parkes et al., 2018).

In-scanner head motion is thought to bias connectivity estimates between two nodes in relation to the distance between the nodes (Ciric et al., 2017; J. D. Power, Barnes, Snyder, Schlaggar, & Petersen, 2012; Theodore D Satterthwaite et al., 2012). This observation is particularly relevant for neurodevelopmental cohorts, where increased in-scanner motion distinguishes patients from controls (Theodore D Satterthwaite et al., 2012). Subject movement may enhance short-distance connections while diminishing long-distance connections (Ciric et al., 2017; J. D. Power et al., 2012; Theodore D Satterthwaite et al., 2012). To assess the residual distance dependence of subject movement post-denoising for two pipelines, we calculated the QC-FC distance dependence (Parkes et al., 2018). We initially employed the centre of mass for each node to determine the Euclidean distance between the nodes $i$ and $j$. Subsequently, we computed the correlation between the distance separating each node pair and the QC-FC correlation of the edge linking those nodes; this correlation served as an estimate of the distance-dependence of motion artifacts.

Figure S2 displays the QC-FC distributions for our primary and alternative pipelines, as well as distance dependence for evaluating residual motion contamination in our sample. All QC-FC values fell within the range of $-0.3 < r < 0.3$, with 95% of the values situated between $-0.15 < r < 0.16$, signifying that residual correlations between FC and head motion were minimal. These values are comparable to previous benchmarking studies on pipeline denoising efficacy, where some of the most effective denoising pipelines exhibited QC-FC values ranging between $-0.45 < r < 0.45$ (Parkes et al., 2018). Similarly, although some QC-FC distance dependence persists, the relationship aligns with earlier findings (Parkes et al., 2018), underscoring the relative success of our denoising methodology.

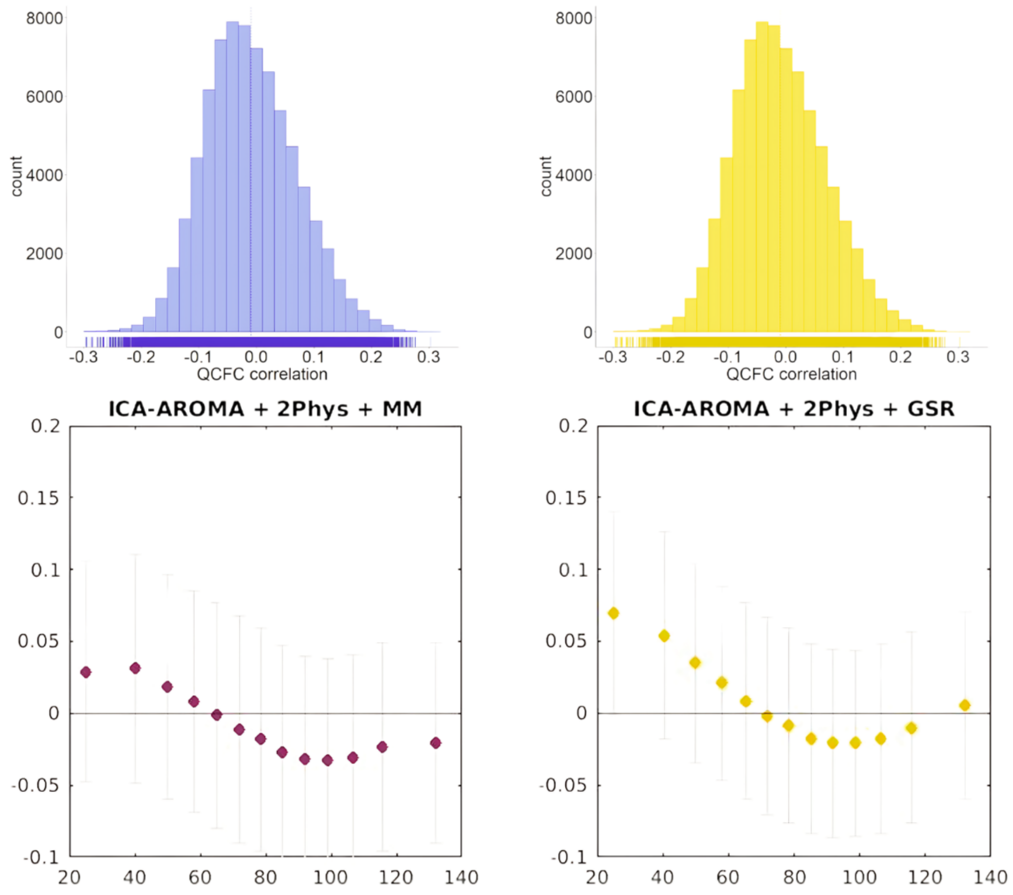

**Figure S2. QC-FC distributions and distance dependence**
The distributions of QC-FC correlations across edges indicate that this dependence is minimal, with both preprocessing pipelines yielding distributions centred near zero. The QC-FC distance dependence plots show the relationship between QC-FC correlation and the spatial distance of each edge. For every bin, the mean QC-FC for the ROI distance bin (circle) and the standard deviation (error bar) of the QC-FC correlation are presented.

**Table S1. Scanning parameters for each scanning site in the LEAP consortium**

| | | | | **EPI BOLD multi-echo imaging sequence** | | | |
|---|---|---|---|---|---|---|---|
| **Site** | Scanner | Field str. | Instruct. | TR /TE1/TE2 / TE3 (ms)/FA (°) | No. of vol. | No. of slices | Voxel size |
| **Cambridge** | Siemens Magnetom Verio | 3 T | Fixation | 2300/ 12/29/46/80 | 266 | 33 | 3.8 x 3.8 x 3.8 |
| **KCL** | GE NA | 3 T | Fixation | 2300/ NA/31/48/90 | 215 | 33 | 3.8 x 3.8 x 3.8 |
| **Mannheim** | Siemens Magnetom TIM Trio | 3 T | Fixation | 2300/ 12/29/46/80 | 215 | 33 | 3.8 x 3.8 x 3.8 |
| **Nijmegen** | Siemens Magnetom Skyra | 3 T | Fixation | 2300/ 12/NA/NA/80 | 266 | 33 | 3.8 x 3.8 x 3.8 |
| **Utrecht** | Philips | 3 T | Fixation | 2300/ 13/31/49/80 | 200 | 33 | 3.75 x 3.75 x 3.75 |

### Table S2. Scanning parameters for each scanning site in the ABIDE 1 cohort

| | | | | EPI BOLD imaging sequence | | | |
|---|---|---|---|---|---|---|---|
| **Site** | Scanner | Field str. | Instruct. | TR /TE (ms)/FA (°) | No. of vol. | No. of slices | Voxel size |
| **Caltech** | Siemens Magnetom Verio | 3 T | Eyes closed | 2000/30/75 | 150 | 34 | 3.5 x 3.5 x 3.5 |
| **CMU** | Siemens Magnetom Verio | 3 T | Eyes closed | 2000/30/73 | 240 | 28 | 3.0 x 3.0 x 3.0 |
| **KKI** | Philips Achieva | 3 T | Fixation | 2500/30/75 | 156 | 47 | 3.0 x 3.0 x 3.0 |
| **Leuven** | Philips | 3 T | Fixation | 1667/33/90 | 250 | 32 | 3.6 x 3.6 x 4.0 |
| **MaxMun** | Siemens Magnetom Verio | 3 T | Fixation | 3000/30/80 | 120/200 | 40 | 3.0 x 3.0 x 3.0 |
| **NYU** | Siemens Magnetom Allegra | 3 T | Fixation | 2000/15/90 | 180 | 33 | 3.0 x 3.0 x 4.0 |
| **Olin** | Siemens Magnetom Allegra | 3 T | Fixation | 1500/27/60 | 210 | 29 | 3.4 x 3.4 x 4.0 |
| **Pitt** | Siemens Magnetom Allegra | 3 T | Eyes closed | 1500/25/70 | 200 | 29 | 3.1 x 3.1 x 4.0 |
| **SBL** | Philips Intera | 3 T | Eyes closed | 2200/30/80 | 200 | 38 | 2.75 x 2.75 x 2.72 |
| **SDSU** | GE MR750 | 3 T | Fixation | 2000/30/90 | 180 | 41 | 3.4 x 3.4 x 3.4 |
| **Stanford** | GE SIGNA | 3 T | Eyes closed | 2000/30/80 | 180 | 29 | 3.1 x 3.1 x 4.5 |
| **Trinity** | Philips Achieva | 3 T | Eyes closed | 2000/28/90 | 150 | 38 | 3.0 x 3.0 x 3.5 |
| **UCLA** | Siemens Magnetom TIM Trio | 3 T | Fixation | 3000/28/90 | 120 | 34 | 3.0 x 3.0 x 4.0 |
| **UM** | GE SIGNA | 3 T | Fixation | 2000/30/90 | 300 | 40 | 3.44 x 3.44 x 3.0 |
| **USM** | Siemens Magnetom TIM Trio | 3 T | Eyes open | 2000/28/90 | 240 | 40 | 3.4 x 3.4 x 3.0 |
| **Yale** | Siemens Magnetom TIM Trio | 3 T | Eyes open | 2000/25/60 | 200 | 34 | 3.4 x 3.4 x 4.0 |

**Table S3. Scanning parameters for each scanning site in the ABIDE 2 cohort**

| | | | | EPI BOLD imaging sequence | | | |
|---|---|---|---|---|---|---|---|
| **Site** | Scanner | Field str. | Instruct. | TR /TE (ms)/FA (°) | No. of vol. | No. of slices | Voxel size |
| **EMC** | GE MR750 | 3 T | Eyes closed | 2000/30/85 | 160 | 37 | 3.6 x 3.6 x 4.0 |
| **ETH** | Philips Achieva | 3 T | Fixation | 2000/25/90 | 210 | 40 | 3.0 x 3.1 x 3.0 |
| **GU** | Siemens Magnetom TIM Trio | 3 T | Eyes open | 2000/30/90 | 154 | 43 | 3.0 x 3.0 x 2.5 |
| **IU** | Siemens Magnetom TIM Trio | 3 T | Eyes open | 813/28/60 | 433 | 42 | 3.4 x 3.4 x 3.4 |
| **KKI** | Philips Achieva | 3 T | Fixation | 2500/30/75 | 156 | 47 | 3.0 x 3.0 x 3.0 |
| **KUL** | Philips | 3 T | Fixation | 2500/30/90 | 162 | 45 | 2.5 x 2.5 x 2.7 |
| **NYU1** | Siemens Magnetom Allegra | 3 T | Fixation | 2000/15/90 | 180 | 33 | 3.0 x 3.0 x 4.0 |
| **NYU2** | Siemens Magnetom Allegra | 3 T | Fixation | 2000/30/82 | 180 | 34 | 3.0 x 3.0 x 3.0 |
| **SDSU** | GE MR750 | 3 T | Fixation | 2000/30/90 | 180 | 41 | 3.4 x 3.4 x 3.4 |
| **Stanford** | GE SIGNA | 3 T | Eyes closed | 2000/30/80 | 180 | 31 | 3.4 x 3.4 x 3.5 |
| **TCD** | Philips Achieva | 3 T | Fixation | 2000/27/90 | 210 | 37 | 3.0 x 3.0 x 3.2 |
| **UCD** | Siemens Magnetom TIM Trio | 3 T | Eyes open | 2000/24/90 | 460 | 36 | 3.5 x 3.5 x 4.0 |
| **USM** | Siemens Magnetom TIM Trio | 3 T | Eyes open | 2000/28/90 | 240 | 40 | 3.4 x 3.4 x 3.0 |
| **Miami** | GE Healthcare | 3 T | Eyes closed | 2000/30/75 | 290 | NA | 3.4 x 3.4 x 3.0 |

### Section 5. Application of Mixture Modeling Normalization

Mixture modeling normalization was implemented under the premise that significant edges are sparse, and the edges approaching zero contribute to within-subject noise. This method functions as soft thresholding, enhancing the signal-to-noise ratio for each individual connectome.

During this process, each correlation value was Fisher-z normalized. Following this, three parametrized distributions were fitted: a central Gaussian distribution to model noise, and two Gamma distributions to model the positive and negative tails, representing the signal. Subsequently, the connectivity values were normalized by subtracting the mean and dividing by the standard deviation of the Gaussian distribution fitted to the noise (Chauvin, Mennes, Llera, Buitelaar, & Beckmann, 2019). For a comprehensive explanation, see (Chauvin et al., 2021; Chauvin, Mennes, Buitelaar, & Beckmann, 2018; Chauvin et al., 2019; Alberto Llera, Huertas, Mir, & Beckmann, 2019; A Llera, Pruim, Wiegerinck, & Beckmann, 2015; A Llera, Vidaurre, Pruim, & Beckmann, 2016; Tyszka, Kennedy, Paul, & Adolphs, 2014).

**Section 6. Removal of scanning-site-specific variance**

To mitigate the impact of scanning site variations, we employed ComBat (Johnson, Li, & Rabinovic, 2007), a harmonization method that uses multivariate linear mixed-effects regression coupled with empirical Bayes. This method is well-documented for its effectiveness in removing site-related artefacts in multi-site resting-state fMRI studies (Yu et al., 2018). In our FC analysis, ComBat was applied while preserving the variance attributed to diagnostic group, age, and sex, while effectively normalizing differences across sites. Further details and validation of this approach on the data used here can be found in (Ilioska et al., 2022).

**Section 7. Normative modeling evaluation**

We report distributions across connections of four metrics to evaluate the performance of the normative models (Figure S3). Explained variance quantifies the Pearson correlation between the predicted and the actual value at each edge. Most values fall within the range of r=0.1 and r=0.2, which is comparable to explained variance reported for functional data (Rutherford et al., 2023). Furthermore, mean standardized log loss (MSLL) measures the difference of the log loss of the test data and the log loss under the predicted distribution, standardized over the test data. Lower MSLL values indicate better model performance. The values in our model are close to 0 which indicates that the model performed well. Finally, skewness and kurtosis of the pseudo z-scores distributions measure how well the model distribution matches the observed distribution (Dinga et al., 2021).The pseudo $z$-scores are quantiles of the observed distribution mapped to $z$-scores of a standard normal distribution (P. K. Dunn & Smyth, 1996). The ideal skewness is 0 whereas the ideal kurtosis for normal distribution is 3, indicating that our model performs well also according to these metrics (Figure S3).

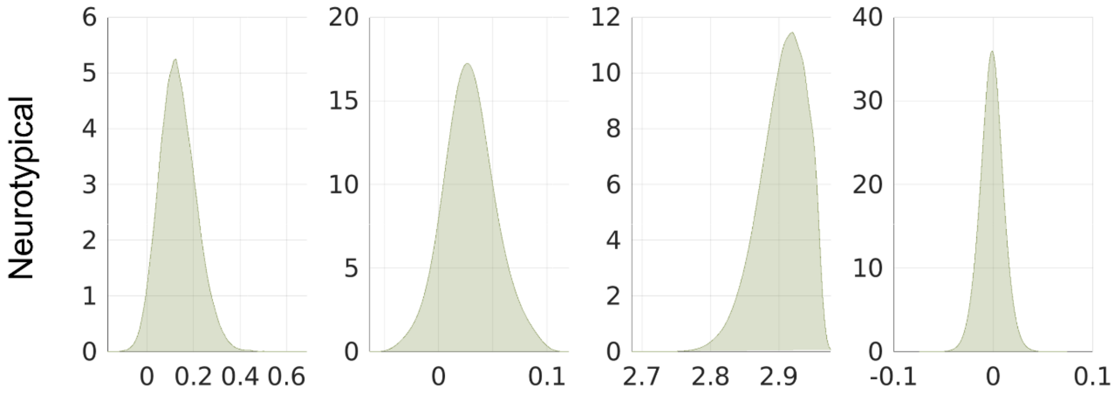

**Figure S3. Distributions of the normative modelling evaluation metrics (columns) for the group of neurotypical individuals.**

Explained variance by the normative model across edges; Mean Standardized Log Loss (MSLL) comparing the model's predicted distribution to the observed data; Kurtosis and skew of the distribution of pseudo-z scores.

**Section 8. Behavioral variables**
Our analysis includes a range of behavioural measures essential for assessing various aspects of autism spectrum disorders (ASD). These include the Autism Diagnostic Observation Schedule (ADOS) calibrated severity scores for social affect, communication, and restrictive and repetitive behaviors, and the Autism Diagnostic Interview-Revised (ADI-R) scores across the social, communication, and restricted and repetitive behaviors domains. Additionally, we utilized the scores from the Social Responsiveness Scale second edition (SRS-2) (Constantino & Gruber, 2012), The Short Sensory Profile (SSP) subscales (McIntosh, Miller, Shyu, & Dunn, 1999) were also included to evaluate sensory processing difficulties, the Autism-Spectrum Quotient (AQ) (Baron-Cohen, Wheelwright, Skinner, Martin, & Clubley, 2001) and full scale IQ assessed using the Wechsler Abbreviated Scales of Intelligence—Second Edition (Wechsler, 1999).

The ADI-R provides insights into an individual's developmental history through a structured interview conducted with parents or caregivers (Rutter, Le Couteur, & Lord, 2003). In contrast, the ADOS assesses current symptoms through a semi-structured interaction between the individual and an examiner (Lord et al., 1999; Reaven, Hepburn, & Ross, 2008). For the purposes of this study, ADOS 2 scores from the ABIDE dataset were converted to calibrated severity scores (Hus, Gotham, & Lord, 2014; Hus & Lord, 2014) to maintain consistency with LEAP score methodologies.

The SRS-2 measures the presence and severity of social difficulties in autism, with assessments based on parent and/or teacher ratings for individuals under 18, and self-reports for those aged 18 and older (Constantino & Gruber, 2012).

The Short Sensory Profile (SSP) is a brief version of Dunn's Sensory Profile, a caregiver questionnaire designed to screen and identify children with sensory processing difficulties (W. Dunn, 1999; McIntosh et al., 1999). It comprises 38 items across seven subscales: Movement Sensitivity, Tactile Sensitivity, Sensation Seeking/Under-responsiveness, Visual/Auditory Sensitivity, Low Energy/Weak, Taste/Smell Sensitivity, and Auditory Filtering. To enhance the interpretability of the results, we inverted the SSP subscale scores such that lower scores now indicate less severe symptoms. To summarize the SSP in two variables, in our analysis we used summary measures of sensory hypo- and hypersensitivity calculated according to (Lefebvre et al., 2023).

AQ (Baron-Cohen et al., 2001) is a continuous measure, administered via self-report or parent-report, designed to quantify the extent of behavioral traits associated with Autism Spectrum Disorder (ASD) in children, adolescents, and adults of average intelligence.

**Section 9. Results for t=1.7 and t=3.1**
Since the choice of threshold could impact the observed results, we repeated our analyses applying the more liberal threshold t=1.7 ($p = 0.001$) and the more conservative threshold of t=3.1 ($p = 0.05$) to examine the overlap of participants across the 3 levels. In line with our original analysis, no significant overlap difference was found on the level of individual connections for the two alternative thresholds. On the level of regions and networks, the results for t=1.7 showed that subjects overlapped more in general (Figure S4 A) compared to our original analysis (Figure 3B) which is expected given the lower threshold. Statistical comparison of the overlaps between autistic and neurotypical individuals revealed a lower abut more wide-spread relative difference for both positive and negative deviations reaching

up to 1,35 in some regions and 2,1 times more overlap in some networks, exhibiting similar pattern to our reported results on both region and network level.

The results for the higher threshold of t=3.1, showed lower overlap across thresholds on both, region and network levels. Including higher but less wide-spread positive deviation difference, on both levels, and negative deviation difference on the level of regions. There was no significant group difference on the network level for negative deviations (Figure S4 B).

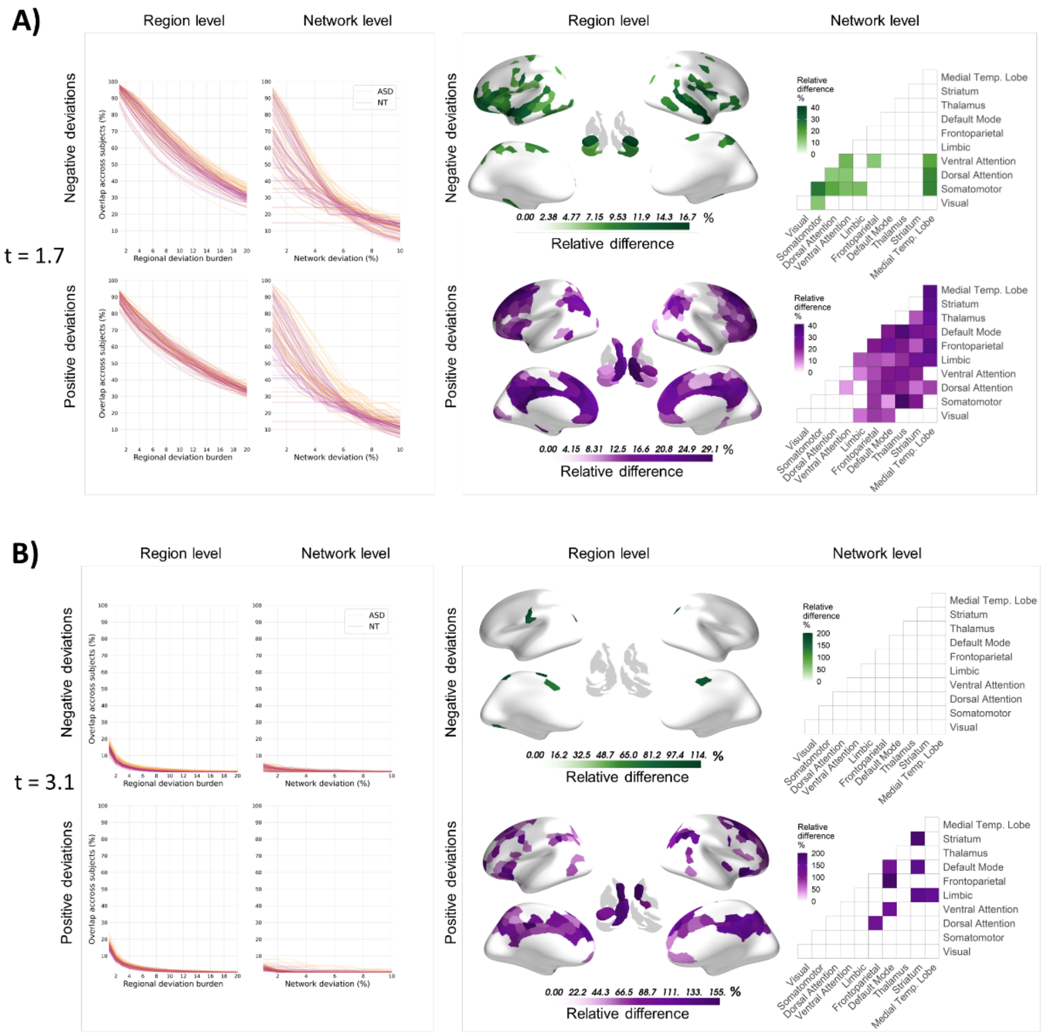

**Figure S4. Overlap of negative and positive functional connectivity (FC) deviations from the normative model and relative difference in overlap between autistic and neurotypical individuals on the level of connections, regions and networks for alternative thresholds.** A) Overlap across thresholds and difference of overlap between groups for extreme deviation threshold t=1.7 corresponding to probability (p=0.001) and B) respectively t=3.1 corresponding to probability (p=0.05). Line plots showing the overlap of subjects on the level of regions and networks. The first column shows the overlap for positive and negative regional deviation burden across 20 thresholds. Each line represents a region in the brain. Yellow color indicates overlap across autistic individuals, whereas red indicates overlap across neurotypical controls. The second column shows the overlap of negative and positive network deviation percent. Here, each line represents a within or between network overlap across participants, where yellow indicates overlap across the group of autistic individuals and red indicates the group of controls. D) Regions and networks showing significantly greater overlap in autistic individuals represented as relative differences. Green color indicates overlap of negative deviations; purple color indicates overlap of positive deviations.

**Section 10. Sensitivity analyses**

In order to test the sensitivity of our results to participant's sex, attention deficit hyperactivity disorder (ADHD) comorbidity and psychotropic medication, we performed our analyses 1) Including only male participants (655 autistic with mean age 16.4 years; 772 neurotypical with mean age 16.3 years; 2) excluding participants with ADHD comorbidity (651 autistic with average age of 16.9 years; 105 female; 1018 neurotypical with average age 15.9 years; 250 female) and 3) excluding all subjects that take psychotropic medication (591 autistic with average age of 16.7 years; 99 female; 1007 neurotypical with average age 15.9 years; 250 female). We report the Pearson correlation of the between group difference result from the sensitivity analyses with the original analysis between-group difference in Table S4, both on the network and region level.

**Table S4. Pearson correlations of sensitivity analyses results with the original analysis result**

| | **Networks** | | **Regions** | |
|---|---|---|---|---|
| | **Negative deviations** | **Positive deviations** | **Negative deviations** | **Positive deviations** |
| **Only males** | 0.92 | 0.95 | 0.8 | 0.9 |
| **No ADHD comorbidity** | 0.99 | 0.98 | 0.8 | 0.93 |
| **Unmedicated** | 0.98 | 0.98 | 0.81 | 0.94 |
